# Evaluating the Effectiveness and Implementation of an Organizational Model Promoting Interprofessional Collaboration in Home Care (RIAP): Protocol for a Multi-Method Study Using the RE-AIM Framework

**DOI:** 10.64898/2026.03.26.26349047

**Authors:** Eleni-Marina Ashikali, Fanny Vallet, Serguei Rouzinov, Hubert Maisonneuve, Catherine Busnel

## Abstract

**Background:** With an aging population and increasing comorbidities, supporting people to remain at home for as long as possible is essential. A major challenge in home care is ensuring coordinated, interprofessional collaboration among all actors involved: patients and their close ones, health and social care professionals, and the broader health and social network. To address this challenge, a new organizational model promoting reinforced interprofessional collaboration, the “Réseau Interprofessionnel Ambulatoire de Proximité” (Local Interprofessional Outpatient Network; RIAP) has recently been developed in the canton of Geneva. The aim of this paper is to present the protocol for a research project evaluating both the effectiveness and implementation of this model.

**Methods:** This is a type-2 hybrid effectiveness implementation study guided by the RE-AIM framework, aiming to evaluate the RIAP organizational model and its implementation, context, and outcomes. This multi-method study will examine the RIAP model under real-world conditions. Routinely collected data on patient outcomes and institutional processes will be used to compare the RIAP model with usual care. Questionnaires will assess patients’ perceptions of continuity of care, professionals’ perceptions of interprofessional collaboration, and stakeholders’ views on the acceptability of the model. Open-ended questions will be included to explore experiences and insights in greater depth, complemented by qualitative data on barriers and facilitators to implementation. Financial indicators will also be analyzed to contextualize the model within the institutional setting.

**Results:** The EFFI-RIAP project began in October 2025 and has a planned duration of 22 months. The project will primarily rely on the reuse of institutional routine data, complemented by questionnaires and qualitative data collection scheduled for September 2026. At the time of submission, study preparation and institutional data processing are ongoing.

**Discussion:** This research will provide insights into the clinical, organizational, and implementation-related effects of the RIAP model compared with usual care. The natural deployment of new RIAP teams will enable investigation of the organizational model under real-world conditions. The combined evaluation of the effectiveness and implementation of RIAP will assess the model’s added value, inform refinement, and identify potential barriers and facilitators relevant to implementation in other teams.

## Background

Staying healthy and living at home for as long as possible is a common goal among adults [1, 2], and many countries have introduced policies on aging in place [3]. Home care services play a central role in achieving this aim, particularly for individuals with complex health needs. In Switzerland, as in many Western countries, home care has expanded and diversified, offering increasingly specialized medical treatments and services [4]. At the same time, demographic aging, rising frailty and chronic illnesses, combined with shorter hospital stays have intensified demand for home care [5]. Yet home care institutions must respond to these growing health and support needs within a context of fragmented care delivery [6], a large number of decentralized health professionals [7], and partial reimbursement of care services [8, 9].

These broader structural pressures are also reflected in clinical practice. Home care serves a heterogenous population-children, adults, and older adults-with a wide range of pathologies and care requirements [10]. Patients often face overlapping medical, socioeconomic, mental health and organizational challenges that create uncertainty and require a collective approach to care [11]. This growing complexity places demands not only on patients and their families, but also on professionals and the organizations in which they work [12]. Such situations, characterized by multiple interrelated problems, health risks, and extensive care needs [12], call for stronger coordination and collaboration among all stakeholders, including health and social care professionals, patients, and their close ones [13–15]. Ensuring quality and safety in home care, therefore, is supported by effective interprofessional collaboration and reinforced continuity and coordination of care.

These requirements indicate that the current organization of home care delivery may warrant reconsideration. Organizational models characterized by multiple parallel professional relationships can limit opportunities for structured and proactive coordination. Home care nurses may collaborate with numerous primary care providers across their caseload, and conversely, physicians may interact with multiple nurses across different teams. Such fragmentation can hinder efficient collaboration, communication, and care planning [16]. In response to these challenges, the RIAP organizational model (Réseau Interprofessionnel Ambulatoire de Proximité; Local Ambulatory Outpatient Network) was developed and implemented in the canton of Geneva (Switzerland). This model aims to foster structured interprofessional partnerships and enhanced coordination.

### Interprofessional Collaboration

Interprofessional collaboration, defined as a “direct collaboration between professionals from different disciplines and professions”, aims to coordinate patient follow-up, ensure continuity of care, address potential issues that exceed the capacity of a single profession, “optimize the quality of care, and increase economic efficiency” [17, 18]. To achieve interprofessional collaboration in primary care, the key elements to be mobilized are trust, mutual respect, the pursuit of common goals [19], interdependence, perceptions and expectations of other healthcare professionals, their skills, their interest in collaborative practice, the definition of their role, and communication [20]. While interprofessional collaboration is widely recognized as essential for promoting quality care [21], it remains challenging to implement in practice, and requires adaptation to the local context [22, 23]. As such, a range of factors must be incorporated into its local operationalization, such as the geographical distance between professionals who often work in different organizations, the type of care, the health status of patients who are often frail, vulnerable, and/or complex, the referral procedures, the rotation of practitioners, and the communication methods [24].

Studies suggest that interprofessional collaboration improves the quality of care by promoting a comprehensive approach that takes into account the diverse needs of patients through the involvement of professionals from different disciplines [25, 26]. Moreover, multimorbid patients seem to report positive experiences with this type of care [27]. In terms of health outcomes, research has suggested that interprofessional care is beneficial for certain patient subgroups, such as those at cardiovascular risk [28], but has also noted a lack of studies examining the effects of interprofessional collaboration on health, particularly in elderly or multimorbid patients [28, 29].

### Continuity of Care and Care Coordination

Continuity of care is defined as “the extent to which a series of discrete healthcare events is experienced by individuals as coherent and interconnected over time, and consistent with their health needs and preferences” [30, p. 8]. It is “a complex and multifaceted concept” [31, p. 3] encompassing several dimensions. These include *informational continuity* referring to the availability and appropriate use of information related to patients’ care events and circumstances; *relational or longitudinal continuity*, defined as “a history of interactions with the same healthcare professional(s) across a series of discrete episodes” [30, p. 17]; *interpersonal continuity*, reflecting patients’ subjective experience of a caring relationship with their healthcare professional; and *management continuity*, described as “the effective collaboration of teams across care boundaries to deliver seamless care” [30, p., 17] and respond to patients’ fluctuating needs [32, 33]. Together, these dimensions highlight that continuity of care relies on sustained relationships, effective information transfer, coordination among providers, and responsiveness to evolving needs [32]. While continuity of care has traditionally been associated with the dyadic physician–patient relationship, it now encompasses a broader caregiver–care recipient relationship in which the patient plays a central role [30].

Care coordination is defined as “a proactive approach that brings together healthcare professionals and service providers to address the needs of service users and ensure they receive integrated, person-centered care across different settings” [30, p. 8]. It includes activities targeting the patient, family, and caregivers; health and social care professionals and services; and activities that bring together patients and professionals [13]. Adequate care coordination is essential for ensuring continuity of care, whereas insufficient coordination can undermine it [34]. To clarify the competencies required to coordinate care effectively in complex situations, a reference framework on the nursing competency of “coordinating care and services for persons with complex needs” has identified its constituent dimensions and observable indicators [13].

Continuity and coordination of care are closely interrelated concepts, grounded in sustained patient-provider relationships, as well as effective communication and cooperation among professionals [35]. “Continuity enables care coordination by creating the conditions and relationships necessary to foster seamless interactions among multiple providers within interdisciplinary teams or across care settings or sectors” [30, p., 12]. In turn, care coordination supports continuity through the management of care transitions, and the organization of roles and responsibilities among professionals involved in patient care [30].

The dimensions of continuity and coordination of care operate at multiple levels, including the individual level (e.g., continuity of services and linkages across different phases of care, from health promotion to rehabilitation), the organizational level (e.g., multidisciplinarity and information sharing), and the system level [36]. Care characteristics that facilitate continuity of care can be grouped into two broad categories: those related to care management (e.g., care planning and coordination) and those related to service delivery (e.g., uninterrupted provision of care). Accordingly, indicators of continuity of care encompass not only traditional quantitative measures, such as expenditures, avoidable hospitalizations, readmissions, and outpatient service use [37], but also qualitative measures, including stakeholders’ perceptions and satisfaction [38]. Interventions that strengthen continuity and coordination of care, particularly when implemented in collaboration with community-based organizations, have been associated with positive outcomes, such as increased patient satisfaction [39], reductions in hospitalizations [31], readmissions, and visits to the emergency department [40], decreased mortality [41], and lower healthcare costs [40]. Moreover, from the professionals’ perspective, continuity of care is perceived as beneficial and has been associated with increased job satisfaction [38].

### The RIAP Organizational Model

The Institution genevoise de maintien à domicile (Geneva Home Care Institution; IMAD) is an autonomous public institution in the Canton of Geneva and a key player in home care. In the early 2000s, IMAD implemented a nurse-led care model, in which a case manager nurse (CMN) serves as the primary point of contact for patients and their families, conducting clinical evaluations, developing care plans with the patient, family, and primary care physician (PCP), coordinating care delivery, and providing clinical services directly. This model seeks to strengthen understanding of patients and their care networks to support the development of common goals. Multidisciplinary teams, including healthcare professionals beyond the CMN (e.g., occupational therapists, community health care assistants), are organized within defined geographical sectors to minimize travel time. Despite this structure, care delivery often remains siloed, requiring CMNs to coordinate with numerous PCPs and other professionals involved in the care of their assigned patients. (Figure 1a).

**Figure 1.**
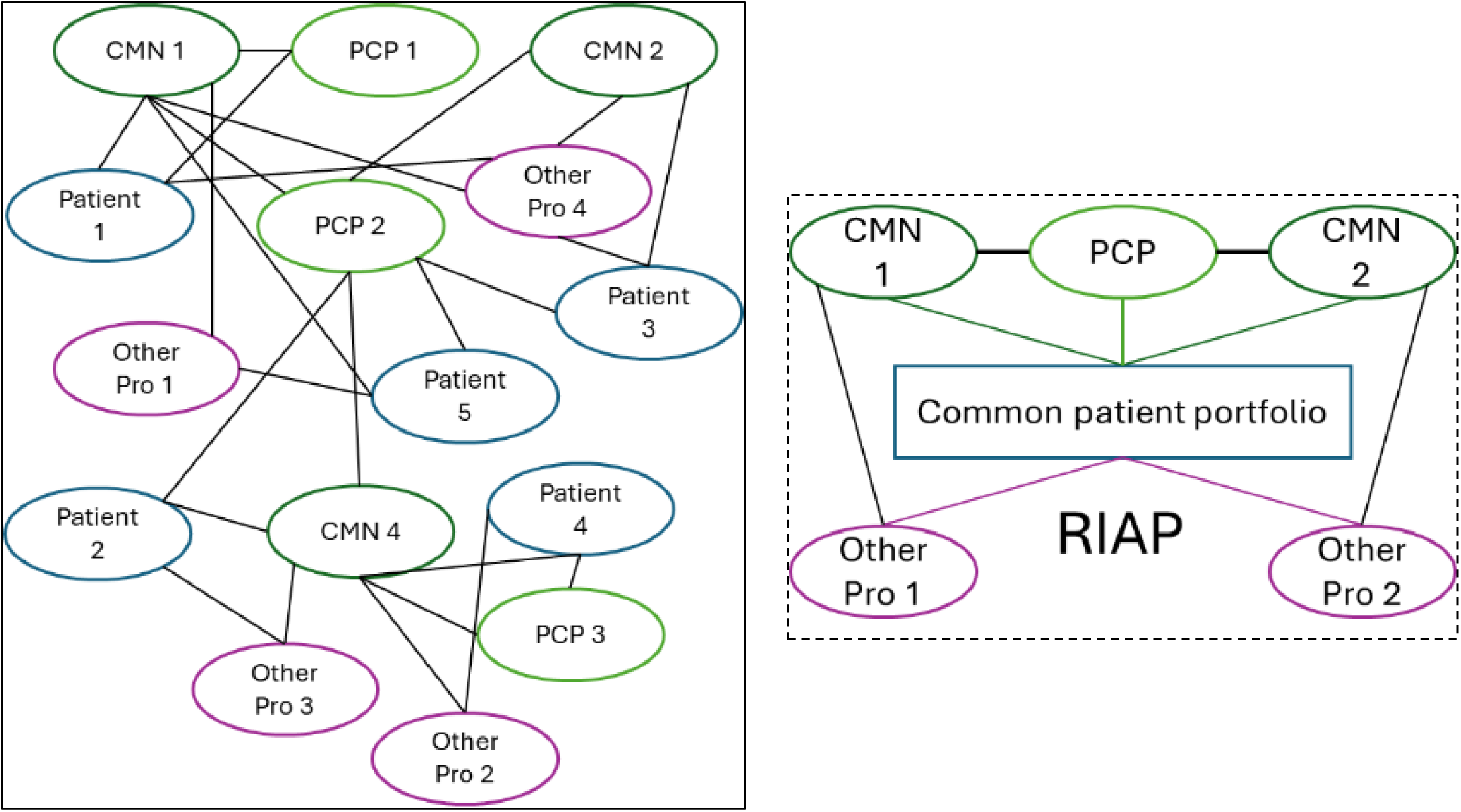
(a) Illustration of the traditional nurse referent model (left). (b) Illustration of a RIAP team with a common patient portfolio (right). Note: PCP = primary care physician; CMN = case manager nurse

In response to these challenges, the RIAP organizational model was introduced within the institution in 2021 [42]. This model puts forward a reinforced interprofessional collaboration using common patient portfolios shared between CMNs and PCPs (Figure 1b). The RIAP model, which was developed according to the principles of the Swiss Academy of Medical Sciences charter [24, 43], aims to optimize home care, particularly in complex situations, by avoiding crisis situations and determining care plan objectives between different professionals, patients, and their close ones. “RIAP is defined as an interprofessional group which aims to provide comprehensive, integrated, and coordinated care by (a) collectively managing a shared patient portfolio while maintaining independent practice, (b) coordinating regularly to evaluate goals established with patients and their caregivers, (c) jointly anticipating possible care actions, and (d) utilizing patients’ intrinsic and extrinsic resources” [24, p. 431]. Following the first implementation, the RIAP model has been, and continues to be, deployed across multiple geographical sites in the canton of Geneva.

### The Present Project: EFFI-RIAP

To guide the evaluation of the RIAP model, the EFFI-RIAP project is framed conceptually using the RE-AIM framework for implementation research [44, 45]. RE-AIM provides a structured approach for examining both the effectiveness of interventions and their implementation in real-world settings, emphasizing factors that influence outcomes on multiple levels, including individual (micro) and institutional (meso) contexts [22]. By highlighting the interplay between patient-, provider-, and system-level factors, this framework supports a comprehensive understanding of how the RIAP model functions, and how it may be adopted, implemented, and maintained in routine home care practice.

## Methods

### Ethical Considerations

The project was submitted to the local ethics committee (Commission Cantonale d’Ethique de la Recherche sur l’être humain; CCER) and was deemed to fall outside of the scope of the Law on Research Involving Human Subjects. A waiver of ethical review was granted on 13.01.2026 (Reference: Req-2025-01715; Req-2025-01716). The waiver was granted on the basis that the project (1) does not aim to obtain generalizable knowledge about human disease, the structure and function of the human body, but primarily collects participants’ perspectives, and (2) constitutes a quality care project evaluating an institutional organizational change. The project will be conducted in accordance with the Declaration of Helsinki and will be carried out in compliance with the Swiss Federal Act on Data Protection.

For routinely collected data generated during the provision of home care services by IMAD, consent is covered by the written service agreement between clients and the institution. This agreement includes a provision permitting the use of de-identified routine care data for scientific research purposes. The ethics committee approved the secondary use of these data without the need for additional written consent.

For project-specific questionnaires, participants, both patients and professionals, will receive comprehensive information about the study, provide written informed consent, and participate on a voluntary basis. They will also be informed of their right to withdraw from the study at any time and without any consequences for their care. All data used within this research project will be de-identified prior to analysis.

The EFFI-RIAP project was registered on the Open Science Framework (DOI: https://doi.org/10.17605/OSF.IO/3HC2R), and a preprint version of this protocol is available on medRxiv (DOI: XXXXX).

### Aims and objectives

The primary objectives of the project are to evaluate the effectiveness and implementation of the RIAP organizational mode. The RE-AIM framework will be used to guide this evaluation, offering a holistic approach to the examination of the model, with findings providing insight not only into its effectiveness and implementation, but also to the people it will reach, its adoption by professionals, and its maintenance through the course of time. The specific research questions (RQs), mapped onto the RE-AIM domains, are presented below, with primary RQs listed first. Figure 2 provides a visual overview of this mapping.

**Figure 2.**
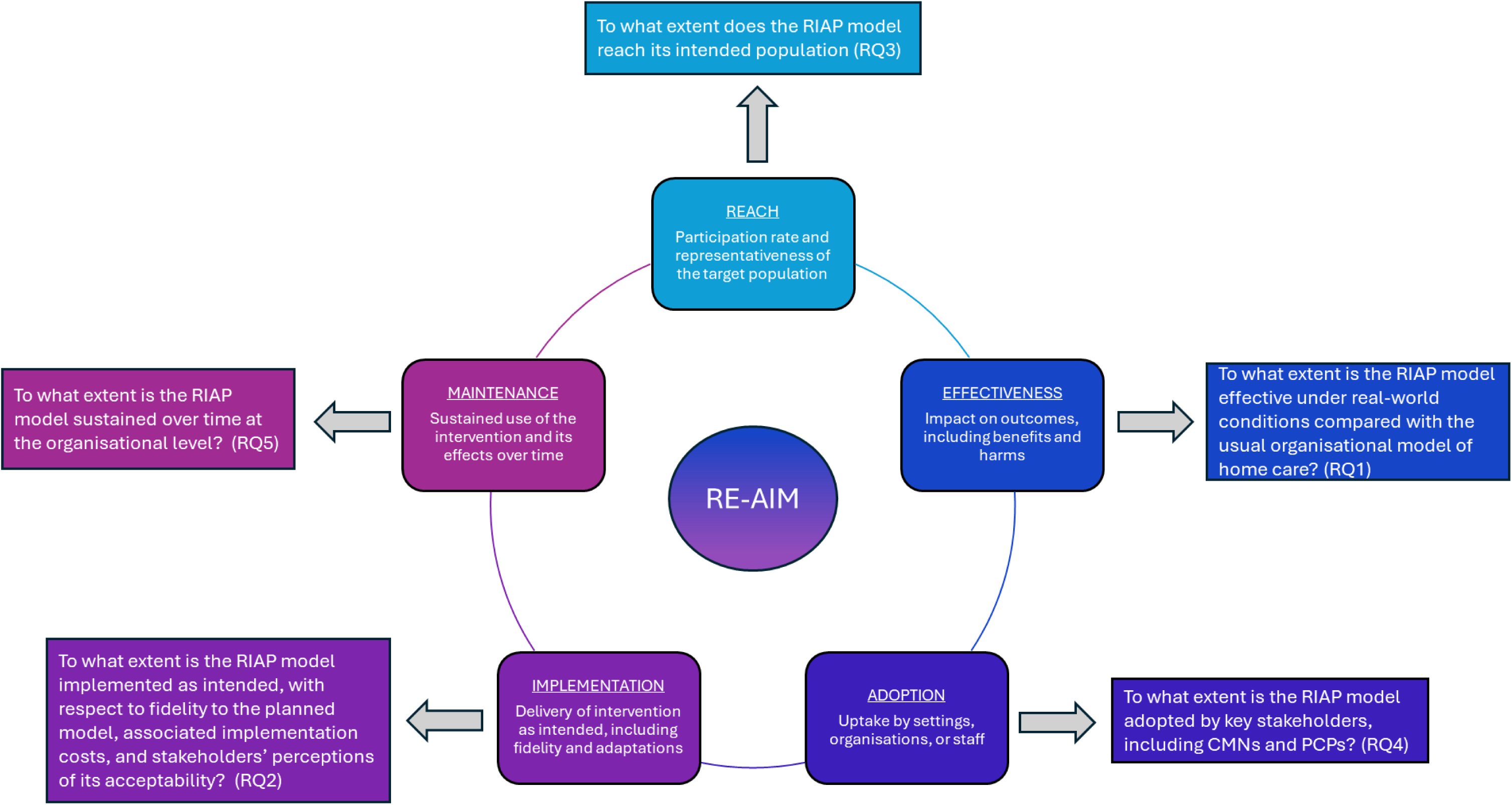
RE-AIM framework domains with corresponding research questions

**RQ1.** To what extent is the RIAP model effective under real-world conditions compared with the usual organizational model of home care? *(Effectiveness)*

**RQ2.** To what extent is the RIAP model implemented as intended, with respect to fidelity to the planned model, associated implementation costs, and stakeholders’ perceptions of its acceptability? *(Implementation)*

**RQ3.** To what extent does the RIAP model reach its intended patient population? *(Reach)*

**RQ4.** To what extent is the RIAP model adopted by key stakeholders, including CMNs and PCPs? *(Adoption)*

**RQ5.** To what extent is the RIAP model sustained over time at the organizational level? *(Maintenance)*

These research questions guide the evaluation of multiple outcomes. Key descriptive outcomes, measured only among RIAP patients or professionals-including patient-reported continuity of care, professional perceptions of interprofessional collaboration, and staff measures, such as work engagement and empowerment-will be described and explored for associations without formal hypothesis testing. For selected outcomes where comparisons between the RIAP model and usual care are possible, such as adverse health events, overall health status, and organizational indicators, including staff turnover-specific hypotheses regarding expected differences or changes over time are pre-specified. These distinctions clarify the scope of the research questions and inform the selection of relevant variables for evaluation.

### Design and setting

The EFFI-RIAP project will employ a multimethod approach to evaluate the effectiveness and the implementation of RIAP model using: (1) A quasi-experimental, design for comparison between RIAP and usual care; (2) Observational and descriptive data about RIAP participants; (3) Qualitative data. The project will be guided by the RE-AIM implementation evaluation framework [44, 45], and reporting will follow the Standards for Reporting Implementation Studies (StaRI) [46] and the REporting of studies Conducted using Observational Routinely-collected health Data (RECORD) [47]. The project will include all teams under the RIAP organizational model as they are naturally deployed across the canton of Geneva (Switzerland), allowing to capture of real-world implementation and performance. Patient-level outcomes will be extracted or collected prior to entry into home care under the new organizational model (pre) whenever feasible, and at a minimum of six months after receiving care under the new model (post). Outcomes will also be compared with data from the institution’s entire patient population (control group). Project-specific questionnaires will be collected in September 2026.

### RE-AIM Framework

Indicator selection for assessing the impact of the RIAP model on patients, healthcare professionals, and the institution is guided by the five domains of the RE-AIM framework [44, 45]. *Reach* refers to the participation of target groups in the program (e.g., patients).

*Effectiveness* captures project outcomes at the end of the project and impacts on key stakeholders (e.g., patients, professionals). *Adoption* reflects the acceptance of the program by relevant stakeholders (e.g., professionals, institution, system). *Implementation* considers adherence to the program protocol, fidelity, and associated costs. Finally, *maintenance* addresses sustainability over time at multiple levels: individual and staff (micro), the organizational (meso), and system or policy (macro). Project outcomes, organized according to these RE-AIM domains, are summarized in Table 1 [please insert below] and reflect the variables selected to assess the impact of the RIAP model on patients, healthcare professionals, and the institution. These domains provide the conceptual structure for the project outcome indicators, which are operationalized in detail in the Measures section.

**Table 1.**
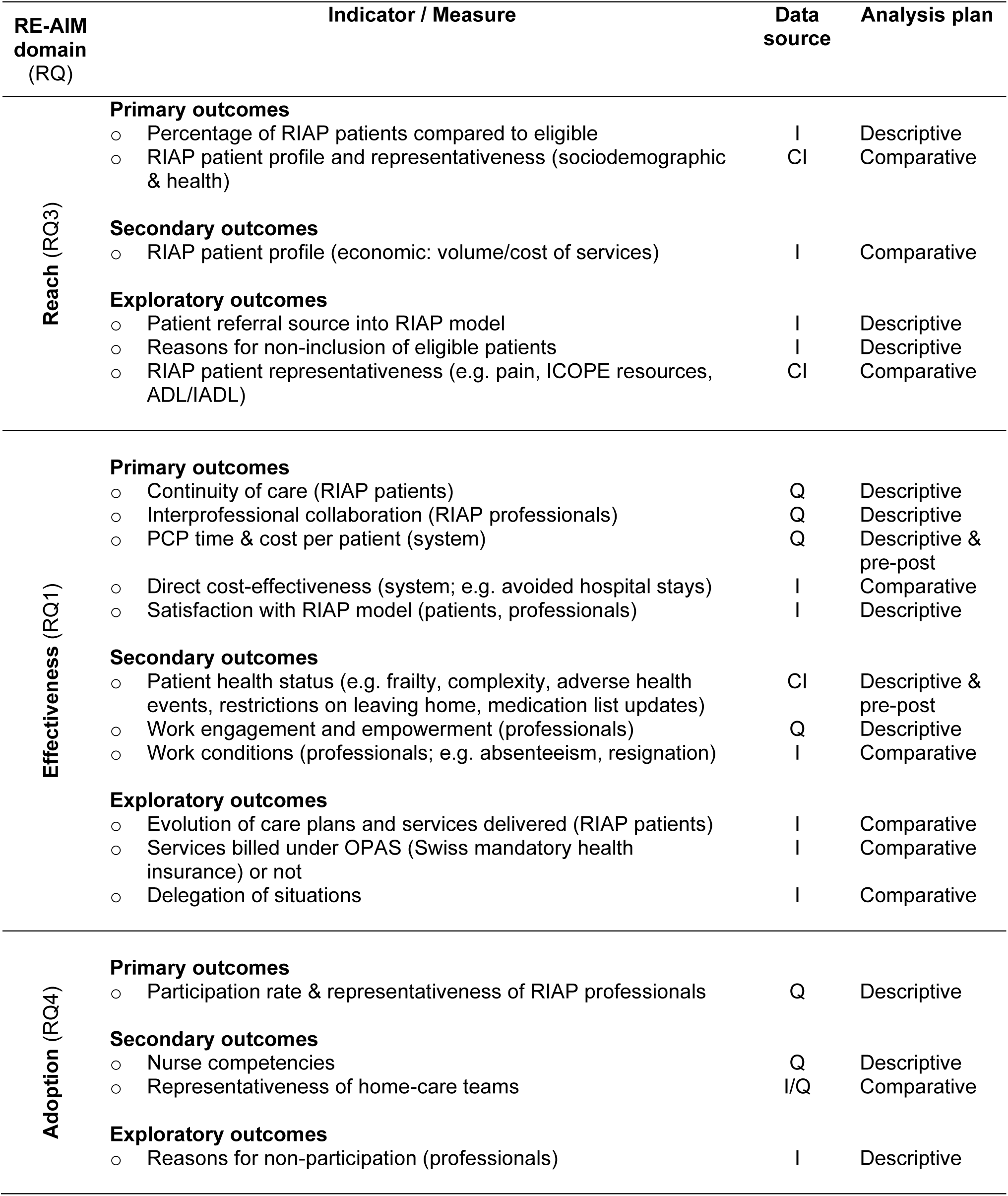
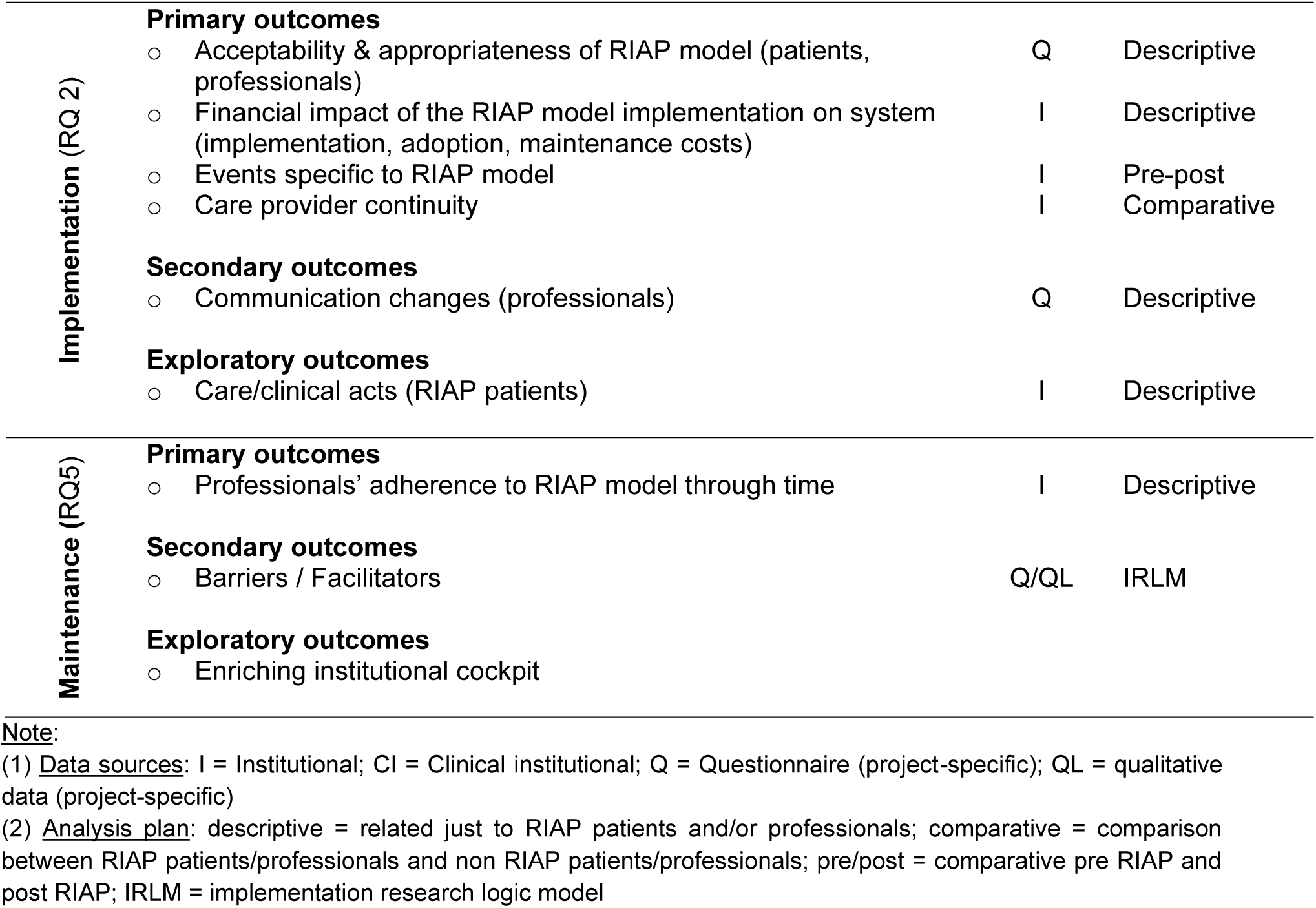
Table outlining the project outcomes, data sources, analysis plans for each RE-AIM domain.

### Participants

The RIAP model is implemented by geographically organized home care teams in collaboration with one or more PCPs to establish a shared patient portfolio. However, only a subset of patients and case nurse managers operate under the RIAP model. The gradual rollout of the model within each team depends on the team’s capacity to manage shared portfolios with PCPs, the willingness of PCPs to adopt the model, and organizational constraints at the team level. Nurse participation is determined in consultation with the team manager.

Participants will be stakeholders related to the RIAP organizational model: patients, CMNs, PCPs, team managers, as well as comparison groups wherever possible (e.g., patients and nurses not within the RIAP model). All participants joined, or will join, the RIAP organizational model in their respective capacities on a voluntary basis. The inclusion criteria are broad in order to best capture the real-world context.

#### Patients

The intervention group will consist of adults (aged ≥ 18 years; expected N = 300), living in the canton of Geneva and receiving home care services by IMAD under the RIAP organizational model. These patients were approached and informed of the new organizational model by their CMN or PCP and consented to receive their care services within it. The comparison group will include all adult patients receiving home care services from IMAD under usual care (N = 7000), with the possibility of stratifying patients into subgroups for specific analyses.

#### Healthcare Professionals

Healthcare professionals will be CMN-PCP dyads (expected N = 20 CMNs and N = 10 PCPs) working under the RIAP organizational model. CMNs are employed within IMAD, whereas PCPs practice externally and participate in the RIAP model for patients they share with the institution. In addition, team managers (expected N = 5) overseeing IMAD teams operating under the RIAP model will be contacted for participation.

### Measures

#### Data Sources

**Qualitative data collection.** Qualitative data will be collected through open-ended questions embedded within the study questionnaires to capture participants’ experiences, perceptions, and contextual factors related to implementation of the organizational model. In addition, participants will be offered the opportunity to take part in follow-up interviews with members of the research team to further explore issues not fully captured in the questionnaires or requiring more in-depth discussion.

Interviews will be conducted using a flexible, semi-structured approach informed by emerging findings and implementation-relevant domains (e.g., acceptability, feasibility, interprofessional collaboration). In addition, facilitators and barriers mentioned by CMNs, PCPs, team managers and decision makers will be systematically documented. The qualitative component is intended to complement quantitative findings by providing explanatory insight into implementation processes.

**Quantitative data extraction and collection.** Quantitative data will be extracted from institutional records routinely collected during standard practice. Institutional records serve multiple functions: clinical documentation (e.g., health needs, care plans), daily care delivery (e.g. monthly volume of services per patient), team management (e.g., number of healthcare professionals involved in a given situation, estimated time to travel from the team office to patient home), quality monitoring (e.g., patient satisfaction) or financial management (e.g., costs). These data are available for all patients receiving home care from the institution (approximately N = 7000), including RIAP patients. Certain institutional data to be extracted will relate exclusively to the EFFI-RIAP project, such as nurse activities within the RIAP model and costs associated with project implementation. Project-specific quantitative data will be collected from patients, CMNs, and PCPs involved in the RIAP model via questionnaires to address specific research questions. Questionnaires not available in French (local language) will be back translated [48] by two members of the research team (EA, SR).

#### Indicators and Measures

To enhance clarity, outcome variables are presented according to each research question (RQ).

**RQ1.** To what extent is the RIAP model effective under real-world conditions compared with the usual organizational model of home care? *(Effectiveness)*

The primary outcomes selected to evaluate the effectiveness of the RIAP model include perceptions of continuity of care (patients), interprofessional collaboration (professionals), satisfaction with the RIAP model (patients and professionals), and cost-effectiveness through avoided hospitalizations and unplanned emergency or urgent care visits (system).

**Patient Experienced Continuity of Care Questionnaire (PECQ**) [49]. The PECQ will be used as a patient-reported experience measure (PREM) of continuity of care. The instrument includes 20 items grouped into four dimensions of continuity of care: information (4 items, α = .86; e.g., “The information I receive from different healthcare professionals is consistent”); relationship (6 items, α = .87; e.g., “I feel safe with the care I receive, regardless of which healthcare professional is looking after me”); management (5 items, α = .88; e.g., “The care staff always help me to coordinate my medical visits”); knowledge (5 items, α = .87; e.g., “I am always involved in planning my care”). Items are rated on a 4-point Likert response scale from 1 *(strongly disagree)* to 4 *(strongly agree)*.

**Interprofessional Collaboration Scale (ICS)** [50]. The ICS will be used to assess interprofessional collaborative relationships. The scale was designed for use by nurses, physicians, and other health professionals working in contemporary acute care settings and uses a 4-point Likert response scale (1 = strongly disagree, 4 = strongly agree). It consists of 13 items grouped into three factors: communication (5 items, e.g., “Physicians and nurses agree on their respective responsibilities”), accommodation (5 items, e.g., “Physicians cooperate with the way we organize nursing care”), and isolation (3 items, e.g., “Physicians generally do not ask for nurses’ opinions”). Versions of the ICS are available for different health professionals (physicians, nurses, and other health professionals), such that ratings by different professionals can be compared. This is particularly interesting in the context of the RIAP model, whereby dyads of healthcare professionals (CMNs, PCPs) work together on common portfolios of patients. Internal consistency in the original version is acceptable (α = .71 to .86), and in translated versions of the scale (German, Italian) the Cronbach’s alpha has been reported at around .92.

**Satisfaction with the RIAP model** will be assessed for both patients and professionals (CMNs and PCPs). For professionals, existing institutional questionnaires will be used, and parallel items will be created for patients to allow for comparisons between their perspectives and those of the professionals.

Secondary outcomes will assess both patient- and nurse-related domains. Patient-related outcomes include the economic profile of RIAP patients, measured in terms of service volume and associated costs, as well as health-related indicators routinely collected in practice, notably the International Resident Assessment Instrument – Home Care (interRAI-HC) [51] and the Complexité Multidimensionnelle de prises en soins Infirmières à Domicile (Multidimensional Complexity in Home Care; COMID) [52]. Nurse-related outcomes within the RIAP model will examine working conditions, including absenteeism, team changes, and resignations, and will be compared with institutional nurses not operating under the RIAP organizational model. Questionnaire-based measures will assess work engagement (Utrecht

Work Engagement Scale; UWES) [53], and psychological empowerment (Psychological Empowerment Instrument; PEI) [54].

**InterRAI-HC** [51]. The interRAI-HC is a standardized, comprehensive assessment tool designed to evaluate the needs, strengths, and health status of community-dwelling older adults receiving home care services. It covers multiple domains, including functional status, cognition, mood, health conditions, social support, and service use. From this tool, several health-related variables will be derived, including self-reported health, adverse health events, pain, individual capacity for activities of daily living, instrumental activities of daily living, frailty, as well as algorithms assessing risks, such as institutionalization (MAPLE) [55]. These subscales have demonstrated good psychometric properties in international and Swiss validation studies, with reported Cronbach’s alpha values generally above .70 and strong evidence of construct and predictive validity (e.g., for outcomes such as functional decline, hospitalization, and mortality). The instrument has been adapted for the Swiss context and has shown reliability and validity in capturing the complexity of frail populations receiving home care.

**COMID** [52]. The COMID is a standardized instrument designed to capture the multidimensional complexity of patient situations encountered by community nurses. It comprises 30 dichotomous items (yes/no) across six domains: medical health factors, mental health circumstances, socioeconomic factors, behavioral factors, instability circumstances, and factors related to the care network and system. Example items include: “The patient’s clinical condition is unstable,” “The patient presents with cognitive impairments,” and “The family environment presents difficulties that complicate care.” A total score will be calculated, with higher scores reflecting greater complexity. The COMID has demonstrated good psychometric properties in Swiss validation studies, with interrater reliability coefficients generally above .70 and strong construct validity, supporting its use in assessing and managing complex home care situations.

**UWES** [53]. The French 9-item abridged version of the UWES scale, validated and shown to possess stronger psychometric properties than the 17-item version [56], will be used to measure RIAP nurses’ engagement with their work. Three 3-item scales cover the domains vigor (e.g. “When I get up in the morning, I feel like going to work”), dedication (e.g. “I am proud of the work that I do”), and absorption (e.g. I am totally immersed in my work”). Items are rated on a 7-point Likert scale from 0 *(never)* to 6 *(always)*. Cronbach’s alphas for the French version of the scale and subscales have been reported between .81 and .92.

**PEI** [54]. This 12-item scale conceptualizes psychological empowerment as a proactive state comprising four cognitions related to work: meaning, competence, self-determination, and impact. Each dimension is measured by three items, such as “The work I do is very meaningful to me” (meaning), “I feel confident about my ability to do my job” (competence), “I have significant autonomy in determining how I do my job” (self-determination), and “I have a great deal of influence in my work group” (impact). Items are rated on a 5-point Likert scale from 1 *(disagree)* to 5 *(strongly agree)*. The French version of the scale [57] has demonstrated good psychometric properties, with Cronbach’s alpha values ranging from .73 to .90.

Exploratory outcomes will examine the evolution of patient health plans and of services delivered, services billed under Swiss mandatory health insurance scheme (OPAS), and the rate of delegation of delegable care acts by CMNs to other professionals.

**RQ2.** To what extent is the RIAP model implemented as intended, with respect to fidelity to the planned model, associated implementation costs, and stakeholders’ perceptions of its acceptability? *(Implementation)*

Primary outcomes at the individual level include the acceptability and appropriateness of the RIAP model (patients and professionals) using the **Acceptability of Intervention Measure (AIM) and Intervention Appropriateness Measure (IAM)** [58]. Each subscale contains four items (e.g. “The RIAP model meets my approval” and “The RIAP model seems suitable”, respectively), rated on a 5-point Likert scale from 1 *(completely disagree)* to 5 *(completely agree)*. Cronbach’s alphas in the original article ranged from .85 to .91.

System-level primary outcomes focus on financial and implementation impacts. These include access costs (i.e. travel time to patient’s home); implementation costs (e.g. managerial planning time, coordination meetings, quality circles, average patient visit time, administrative time, nurse workload rate); adoption costs (e.g. time and cost of training; set-up costs); maintenance costs (e.g. turnover, absenteeism). Fidelity to the organizational model will be assessed through documentation of events specific to participation in the RIAP organizational model (e.g. meetings with the PCP or updating of records), as well as through coordination-related care acts (e.g. coordination, assessments). Finally, care provider continuity, that is, the number of different professionals involved in a patient’s care, will also be tracked and compared to patients under the usual care organizational model.

Secondary outcomes include communication changes between CMNs and PCPs, as reported by professional groups, serving as a measure of the implementation of the RIAP model. Exploratory outcomes will encompass care and clinical acts provided to RIAP patients.

**RQ3.** To what extent does the RIAP model reach its intended patient population? *(Reach)* Primary outcomes for the Reach dimension include the participation rate of patients, defined as the proportion of patients currently receiving care under the RIAP model relative to the number of eligible patients to whom the transition to the new organizational model was proposed. Representativeness will be assessed by comparing the sociodemographic characteristics and health profiles (e.g. frailty, complexity) of participating patients with those of the institution’s overall patient population.

Secondary patient-related outcomes include the economic profile of patients under the RIAP model, assessed in terms of service utilization volume and associated costs (volume/cost of services).

Exploratory outcomes encompass patients’ referral sources into the RIAP model, and reasons why eligible patients were not included,

**RQ4.** To what extent is the RIAP model adopted by key stakeholders, including CMNs and PCPs? *(Adoption)*

Primary outcomes for professionals include participation rate and representativeness. Participation will be assessed based on the number of CMNs and PCPs involved in the RIAP model, while representativeness will be examined using their sociodemographic characteristics and the overall number of professionals engaged in the model.

Secondary outcomes include assessment of nurse competencies and the representativeness of home care teams operating within the RIAP model, which will be compared with teams that have not yet integrated it. **Nurse competencies** will be measured using a questionnaire which was created on the basis of a recently developed framework identifying the core competencies required for coordinating nurses working with persons with complex needs [13]. The framework identifies four dimensions: 1. Work with the individual and their close ones throughout the care process; 2. Establish and maintain continuity of care and services between the individual, health and social care professionals and services; 3. Exercise collaborative leadership within intra- and interprofessional teams; 4. Communicate effectively with the individual, their close ones, and intra-interprofessional teams. These dimensions are operationalized with key indicators, which have been used to derive questionnaire items to explore coordinator nurses’ continued education needs [59]. A total of 16 items reflecting the four dimensions were developed (e.g. “I develop, implement, and adjust an individualized care plan in partnership with the patient and their close ones”).

Exploratory outcomes for this dimension will be the reasons for non-participation of eligible professionals.

**RQ5.** To what extent is the RIAP model sustained over time at the organizational level? *(Maintenance)*

The uptake and sustained use of the model by professionals will be assessed by the number of CMNs and PCPs expressing interest in joining the RIAP model, as well as the number who choose to leave the model and return to the usual organizational structure. Barriers and facilitators will be explored across teams using an Implementation Research Logic Model (IRLM) [60]. Exploratory outcomes will include the enrichments to the institutional cockpit.

### Statistical Analysis

The project will employ a multi-method approach. The quantitative component will rely on a real-world evidence framework, using a pragmatic and adaptive methodology consistent with principles of implementation science. The analytic strategy is designed to capture both the complexity of the RIAP intervention and the diversity of outcomes relevant to its deployment in routine practice. Analyses will include a combination of descriptive and comparative methods aligned with the project objectives.

Descriptive analyses will summarize the characteristics of the population reached by RIAP (e.g., sociodemographic profile, clinical characteristics, types and frequency of interventions, providers involved, timing and sequence of actions). Descriptive statistics will include measures of central tendency, dispersion, and proportions as appropriate.

Comparative analyses will assess differences between individuals within the RIAP model and those who are not. Depending on data distribution and variable types, comparisons will use chi–square tests, t–tests or non–parametric equivalents, and regression models adjusted for potential confounders. When relevant, effect sizes and confidence intervals will be reported.

The study will use a controlled design comparing RIAP and usual care, with before–after assessments conducted whenever feasible, allowing evaluation of changes over time while accounting for a comparison group not exposed to RIAP. Analyses will primarily rely on controlled comparisons between RIAP and usual care; pre–post analyses within groups will be conducted as a complementary approach when feasible. Depending on data availability, this may include (1) difference–in–differences analyses to isolate the effect of the RIAP intervention from underlying temporal trends, (2) multivariable regression models to adjust for case–mix differences and potential confounding factors and (3) sensitivity analyses reflecting the adaptive nature of the methodology (e.g., varying inclusion criteria, subgroup analyses).

In line with implementation science, quantitative findings will be complemented by qualitative analyses aimed at identifying mechanisms of action, contextual influences, facilitators and barriers to implementation, and potential effects not captured by quantitative measures. Qualitative data will be analyzed using an inductive thematic analysis approach. This integration supports a comprehensive understanding of the RIAP model as a complex intervention.

#### Sample size and power considerations

This study includes all eligible patients receiving care under the RIAP model during the study period, with approximately 300 patients expected to contribute data. As recruitment is determined by routine service activity, the sample size is pragmatically fixed rather than driven by conventional a priori power calculations.

In line with recommendations for pragmatic evaluations and real-world implementation studies, the primary focus is placed on precision of effect estimates rather than hypothesis-testing power. Accordingly, confidence intervals will be reported systematically to support transparent interpretation of the magnitude and uncertainty of observed effects.

Once preliminary data are available, exploratory post-hoc assessments (e.g., variance estimates, detectable effect sizes) may be conducted to contextualize the robustness of findings. Where relevant, analyses will account for clustering at the professional or team level and repeated measures, which may influence effective sample size.

### Data Management

The institution’s Information Technology (IT) Department will extract the requested data collected during routine care. At this stage, an encrypted patient identifier will be assigned, data enabling patient identification removed (e.g. first and last name, insurance information), and full date of birth converted to just year of birth. The coded data set will then be securely transmitted to the research team.

The research team, composed of four people, will only have access to the coded data and will at no time be able to identify individual patients or access any other personal identifying information. Extracted data will be stored on secure institutional servers, whereas paper questionnaires will be stored in a locked cabinet accessible only to the research team.

### Dissemination Strategy

No restrictions on publication have been imposed by the funding institution. The research team retains full authority to publish and disseminate findings in the most relevant outlets, regardless of the nature of the results. Scientific, professional, and institutional publications are planned. Findings will also be communicated to patients, caregivers, and the public through newsletters, presentations, workshops, or other appropriate channels to ensure a continued strong partnership.

## Results

The EFFI-RIAP project began in October 2025, with the establishment of the research team, which subsequently developed the study design, methodology, and operational definition of the study variables. The total project duration is planned for 22 months. The project will primarily rely on the reuse of institutional routine data, complemented by project-specific questionnaires and qualitative data collection scheduled for September 2026. At the time of submission of this protocol, the preparatory phase of the study is ongoing, including data extraction, data quality checks, and preparation of the institutional dataset.

## Discussion

This research project aims to evaluate both the effectiveness and implementation of RIAP, a novel organizational model in home care aiming to reinforce interprofessional collaboration and continuity of care, currently deployed in the canton of Geneva, Switzerland.

Guided by the RE-AIM implementation science framework, the project will adopt a comprehensive approach, generating insights not only into the model’s effectiveness and fidelity/cost of implementation, but also its reach, adoption by professionals, and long-term sustainability. A multidimensional evaluation of this kind, capturing both individual and organizational-level outcomes, is particularly important for interventions embedded in complex healthcare systems, as organizational processes and contextual conditions may critically influence both implementation and observed effects [22, 23]. By using primarily routine data, the project will provide real-world evidence on the RIAP model while also establishing a foundation for sustainable, long-term evaluation and monitoring. The real-world setting enhances external validity [61], providing insights that are directly applicable to current home care practice.

The examination of the model under real-world conditions necessarily entails a methodology that is flexible and responsive to contextual factors. This is consistent with the pragmatic orientation of the RE-AIM framework and its emphasis on applicability in routine practice, rather than experimental control. While the project will include pre/post comparisons for selected variables, as well as a comparison group, the analyses will remain observational in nature, which may limit causal inference. Furthermore, as the project will be carried out within a single canton, the findings will have limited generalizability to other regions or healthcare contexts with different organizational structures or population characteristics.

Despite these limitations, the project is expected to generate important contributions. Findings will inform policymakers and home care institutions about the implementation processes, effectiveness, and sustainability of the RIAP model, offering guidance for potential scaling or adaptation. Moreover, identifying facilitators and barriers to implementation can support targeted strategies to enhance adoption and strengthen the model’s effectiveness within routine practice [60]. More broadly, the project will provide insights on interprofessional collaboration and continuity of care, which in previous research, have been associated with improved patient outcomes [31, 40, 41], patient and professional satisfaction [38, 39], and lower healthcare costs [40]. Finally, the examination of patients’ health status may contribute to addressing an existing research gap and support a more nuanced understanding of the potential effects of interprofessional collaboration on multimorbid, complex patients [28, 29].

The EFFI-RIAP project may also provide a foundation for future research. Longer-term follow-up could examine the persistence of patient and organizational outcomes over time, while replication in other settings would provide evidence on the RIAP model’s generalizability. Comparative studies with different organizational models could further strengthen the evidence base for home care innovations and guide implementation strategies across different healthcare systems.

## List of Abbreviations

AIM: Acceptability of Intervention Measure
CMN: Case manager nurse
COMID: Complexité multidimensionnelle pour la pratique infirmière à domicile IAM Intervention Appropriatives Measure
ICS: Interprofessional Collaboration Scale
IMAD: Institution genevoise de maintien à domicile (Geneva Home Care Institution)
interRAI-HC: International Resident Assessment Instrument Home Care PCP Primary care physician
PECQ: Patient Experienced Continuity of Care Questionnaire
PEI: Psychological Empowerment Instrument
PREM: Patient-reported experience measure
RE-AIM: Acronym for Reach, Effectiveness, Adoption, Implementation, Maintenance
RIAP: Réseau interprofessionnel ambulatoire de proximité (Local interprofessional outpatient network)
UWES: Utrecht Work Engagement Scale

## Declarations

### Availability of data and materials

N/A

### AI disclosure

AI-based tools were used in certain parts of the manuscript to refine and clarify the language of text written by the Authors. All content was reviewed and approved by the Authors, who take full responsibility for the manuscript.

## Authors’ contributions

CB, FV, EA and HM contributed to the project conception, design, and methodology. FV and SR developed the data analysis plan; SR drafted the statistical analysis section in this manuscript. The first full draft of the manuscript was written by EA. All Authors read, commented, and approved the final manuscript. CB is the Principal Investigator (PI) of the EFFI-RIAP project.

## Data Availability

Institutional data used in this study are subject to confidentiality restrictions and cannot be shared. Project-specific anonymized data are available from the corresponding author upon reasonable request.

## References

1. Grazia Bedin, M., Droz Mendelzweig, M., Dellepiane, M., & Sobrino Piazza, J.,, Vivre à domicile le plus longtemps possible. Etude sur les logements protégés mandatée par le Canton de Vaud. Rapport d’étude. . 2021, La Source. Institut et Haute Ecole de la Santé.

2. Binette, J.V., K.,, Home and Community Preferences: A National Survey of Adults Age 18-Plus. . 2018: Washington, DC.

3. United Nations Economic Commission for Europe, Policy Brief on Ageing No. 27: Ageing in place: A key policy priority. 2021, UNECE: Geneva.

4. Möckli, N., et al., Care coordination in homecare and its relationship with quality of care: A national multicenter cross-sectional study. International Journal of Nursing Studies, 2023. 145: p. 104544.

5. World Health Organization, The growing need for home health care for the elderly: Home health care for the elderly as an integral part of primary health care services. 2015.

6. Kern, L.M., J.P.W. Bynum, and H.A. Pincus, Care Fragmentation, Care Continuity, and Care Coordination—How They Differ and Why It Matters. JAMA Internal Medicine, 2024. 184(3): p. 236–237.

7. Bakewell, F., Medical silos, social identity, and duty of care: A call for health leaders to improve transitions of care. Healthcare Management Forum, 2025. 38(2): p. 148–151.

8. Gatome-Munyua, A., et al., Reducing fragmentation of primary healthcare financing for more equitable, people-centred primary healthcare. BMJ Global Health, 2025. 10(1): p. e015088.

9. Grandchamp, C.M., S., Revue Médicale Suisse : Système de santé suisse : une machine boulimique. Revue Médicale Suisse, 2024. 20(867): p. 666–671.

10. National Research Council (US) Committee on the Role of Human Factors in Home Health Care., The Role of Human Factors in Home Health Care: Workshop Summary. 2010: Washington (DC): National Academies Press (US).

11. Contandriopoulos, A.-P., et al., *Intégration des soins: Dimensions et mise en œuvre.* Ruptures, Revue Transdisciplinaire en Santé, 2000. 8.

12. Busnel, C., C. Ludwig, and M.G. Da Rocha Rodrigues, La complexité dans la pratique infirmière : vers un nouveau cadre conceptuel dans les soins infirmiers. Recherche en soins infirmiers, 2020. **N° 140**(1): p. 7–16.

13. Karam, M., et al., Nursing Care Coordination for Patients with Complex Needs in Primary Healthcare: A Scoping Review. International Journal of Integrated Care, 2021.

14. Pomey, M.-P., et al., Le «Montreal model» : enjeux du partenariat relationnel entre patients et professionnels de la santé. Santé Publique, 2015. S1(HS): p. 41–50.

15. Gerber, M., Kraft, E., & Bosshard, C., La collaboration interprofessionnelle sous l’angle de la qualité. Bulletin des Médecins Suisses, 2018. 99(44): p. 1524–1529.

16. Nicolet, A., et al., Preferences of older adults for healthcare models designed to improve care coordination: Evidence from Western Switzerland. Health Policy, 2023. 132: p. 104819.

17. Busnel, C., Bridier-Boloré, A., Marjollet, L., & Perrier-Gros-Claude, O.,, La complexité des prises en soins à domicile. Guide pour les professionnels de l’aide et des soins à domicile. 2020, Institution genevoise de maintien à domicile.

18. Office fédéral de la santé publique (OFSP), Programme *de promotion interprofessionnalité dans le domaine de la santé 2017-*2020. 2017.

19. Corser, W.D., A conceptual model of collaborative nurse-physician interactions: the management of traditional influences and personal tendencies. Sch Inq Nurs Pract, 1998. 12(4): p. 325–41; discussion 343-6.

20. Bardet, J.-D., et al., Physicians and community pharmacists collaboration in primary care: A review of specific models. Research in Social and Administrative Pharmacy, 2015. 11(5): p. 602–622.

21. World Health Organization, Framework for Action on Interprofessional Education & Collaborative Practice. 2010: Geneva, Switzerland.

22. Pfadenhauer, L.M., et al., Making sense of complexity in context and implementation: the Context and Implementation of Complex Interventions (CICI) framework. Implement Sci, 2017. 12(1): p. 21.

23. Skivington, K., et al., A new framework for developing and evaluating complex interventions: update of Medical Research Council guidance. BMJ, 2021. 374: p. n2061.

24. Perrier-Gros-Claude, O., Busnel, C., Vallet, F., & Sommer, J., Collaboration dans les soins à domicile : Réseau Interprofessionnel Ambulatoire de Proximité. Revue Medicale Suisse, 2023. 19: p. 430–433.

25. Reeves, S., et al., Interprofessional collaboration to improve professional practice and healthcare outcomes. Cochrane Database Syst Rev, 2017. 6(6): p. CD000072.

26. Xyrichis, A. and K. Lowton, What fosters or prevents interprofessional teamworking in primary and community care? A literature review. International Journal of Nursing Studies, 2008. 45(1): p. 140–153.

27. Davidson, A.R., et al., What do patients experience? Interprofessional collaborative practice for chronic conditions in primary care: an integrative review. BMC Prim Care, 2022. 23(1): p. 8.

28. Bouton, C., et al., Interprofessional collaboration in primary care: what effect on patient health? A systematic literature review. BMC Prim Care, 2023. 24(1): p. 253.

29. Lutfiyya, M.N., et al., The state of the science of interprofessional collaborative practice: A scoping review of the patient health-related outcomes based literature published between 2010 and 2018. PLoS One, 2019. 14(6): p. e0218578.

30. World Health Organization, Continuity and coordination of care: a practice brief to support implementation of the WHO Framework on integrated people-centred health services. 2018: Geneva.

31. Barker, I., A. Steventon, and S.R. Deeny, Association between continuity of care in general practice and hospital admissions for ambulatory care sensitive conditions: cross sectional study of routinely collected, person level data. BMJ, 2017. 356: p. j84.

32. Djukanovic, I., et al., The meaning of continuity of care from the perspective of older people with complex care needs-A scoping review. Geriatr Nurs, 2024. 55: p. 354–361.

33. Haggerty, J.L., et al., Continuity of care: a multidisciplinary review. BMJ, 2003. 327(7425): p. 1219–1221.

34. Aued, G.K., et al., Liaison nurse activities at hospital discharge: a strategy for continuity of care. Rev Lat Am Enfermagem, 2019. 27: p. e3162.

35. Uijen, A.A., et al., How unique is continuity of care? A review of continuity and related concepts. Family Practice, 2011. 29(3): p. 264–271.

36. Khatri, R., et al., Continuity and care coordination of primary health care: a scoping review. BMC Health Serv Res, 2023. 23(1): p. 750.

37. Nicolet, A., et al., Association between continuity of care (COC), healthcare use and costs: what can we learn from claims data? A rapid review. BMC Health Serv Res, 2022. 22(1): p. 658.

38. Reig-Garcia, G., et al., Evaluation and perceptions of a nursing discharge plan among nurses from different healthcare settings in Spain. BMC Health Serv Res, 2022. 22(1): p. 710.

39. Saultz, J.W. and W. Albedaiwi, Interpersonal continuity of care and patient satisfaction: a critical review. Ann Fam Med, 2004. 2(5): p. 445–51.

40. Berkowitz, S.A., et al., Association of a Care Coordination Model With Health Care Costs and Utilization: The Johns Hopkins Community Health Partnership (J-CHiP). JAMA Netw Open, 2018. 1(7): p. e184273.

41. Gray, B., The Cynefin framework: applying an understanding of complexity to medicine. J Prim Health Care, 2017. 9(4): p. 258–261.

42. Perrier-Gros-Claude, O., *De l’interdisciplinarité vers l’interprofessionnalité : une évolution évidente et efficiente dans les enjeux du maintien à domicile à GenèveCréation d’un premier réseau interprofessionnel ambulatoire de proximité (RIAP) : étude de faisabilité à visée d’une implémentation locale*, in *Geneva school of economics and management*, Faculté de médecine. 2022, UNIGE: Genève. p. 108.

43. Académie Suisse des Sciences Médicales, Charte 2.0 La collaboration interprofessionnelle dans le système de santé. 2020.

44. Glasgow, R.E., Vogt, T. M., & Boles, S. M., Evaluating the public health impact of health promotion interventions: the RE-AIM framework. American Journal of Public Health, 1999. 89(9): p. 1322–1327.

45. Glasgow, R.E., Harden, S. M., Gaglio, B., Rabin, B., Smith, M. L., Porter, G. C., Ory, M. G., & Estabrooks, P. A., RE-AIM Planning and Evaluation Framework: Adapting to New Science and Practice With a 20-Year Review Frontiers in Public Health, 2019.

46. Pinnock, H., et al., Standards for Reporting Implementation Studies (StaRI): explanation and elaboration document. BMJ Open, 2017. 7(4): p. e013318.

47. Benchimol, E.I., et al., The REporting of studies Conducted using Observational Routinely-collected health Data (RECORD) statement. PLoS Med, 2015. 12(10): p. e1001885.

48. Brislin, R.W., Back-Translation for Cross-Cultural Research. Journal of Cross-Cultural Psychology, 1970. 1(3): p. 185–216.

49. Ljungholm, L., et al., Measuring patients’ experiences of continuity of care in a primary care context-Development and evaluation of a patient-reported experience measure. J Adv Nurs, 2024. 80(1): p. 387–398.

50. Kenaszchuk, C., et al., Validity and reliability of a multiple-group measurement scale for interprofessional collaboration. BMC Health Serv Res, 2010. 10: p. 83.

51. Morris, J.N., et al., InterRAI Home Care Suisse (interRAI HCSuisse) Instrument d’évaluation, 9. 4, Edition Française Pour la Suisse. 2019: interRAI.

52. Busnel, C., L. Marjollet, and O. Perrier-Gros-Claude, Complexité des prises en soins à domicile : développement d’un outil d’évaluation infirmier et résultat d’une étude d’acceptabilité. Revue Francophone Internationale de Recherche Infirmière, 2018. 4(2): p. 116–123.

53. Schaufeli, W.B., A., UWES Utrecht Work Engagement Scale Preliminary Manual. 2004 Occupational Health Psychology Unit Utrecht University.

54. Spreitzer, G.M., Psychological Empowerment in the Workplace: Dimensions, Measurement, and Validation. The Academy of Management Journal, 1995. 38(5): p. 1442–1465.

55. Hirdes, J.P., J.W. Poss, and N. Curtin-Telegdi, The Method for Assigning Priority Levels (MAPLe): a new decision-support system for allocating home care resources. BMC Med, 2008. 6: p. 9.

56. Zecca, G., et al., Validation of the French Utrecht Work Engagement Scale and its relationship with personality traits and impulsivity. European Review of Applied Psychology, 2015. 65(1): p. 19–28.

57. Boudrias, J.-S., et al., Habilitation Psychologique. Swiss Journal of Psychology, 2010. 69(3): p. 147–159.

58. Weiner, B.J., et al., Psychometric assessment of three newly developed implementation outcome measures. Implementation Science, 2017. 12(1): p. 108.

59. Karam, M., et al., Training Needs Analysis of Primary Care Nurses in Care Coordination: Protocol for a Mixed-Methods Study. Science of Nursing and Health Practices, 2026.

60. Smith, J.D., D.H. Li, and M.R. Rafferty, The Implementation Research Logic Model: a method for planning, executing, reporting, and synthesizing implementation projects. Implement Sci, 2020. 15(1): p. 84.

61. Devane, D., et al., Beyond the binary: integrating “real-world evidence” with randomized trials in contemporary health care. J Clin Epidemiol, 2025. 184: p. 111821.

